# Operating characteristics of analysis methods for clinical trials in viral respiratory disease: A simulation study protocol

**DOI:** 10.64898/2026.06.24.26356481

**Authors:** Johannes M. Schwenke, Felix Herkner, Mutamba Tonton Kayembe, Inge C. Olsen, Matthias Briel, Franz König

**Affiliations:** CLEAR Methods Center, Division of Clinical Epidemiology, University and University Hospital of Basel, Basel, Switzerland; Center for Medical Data Science, Medical University of Vienna, Vienna, Austria; Oslo Center for Biostatistics and Epidemiology, Department of Research Support for Clinical Trials, Oslo University Hospital, Oslo, Norway

## Abstract

Acute viral respiratory infections (ARVIs) are a major cause of hospitalization and death worldwide, yet randomized clinical trials in this setting face substantial challenges in selecting efficient and clinically meaningful primary endpoints. Mortality is often too infrequent to serve as a feasible primary endpoint. Several alternative approaches have been proposed, including ordinal scales, time-to-event endpoints, recovery-based composite outcomes, and longitudinal ordinal models. However, their comparative operating characteristics under realistic ARVI disease courses remain insufficiently understood. We describe a simulation study to compare the type I error and power of commonly used and recently proposed endpoints and analysis strategies for two-arm randomized trials in hospitalized participants with ARVIs. Data will be generated under several mechanisms designed to mimic plausible participant trajectories, including a latent Brownian motion process, a first-order ordinal Markov process, a latent recurrent-event process with frailty, and resampling from individual participant data from the ACTT-2 trial. Simulated outcomes will use 4-, 6-, and 8-level ordinal severity scales and will reflect moderately and severely ill populations, follow-up horizons of 28 or 60 days, varying treatment effects, and sample sizes. Methods to be compared include Markov ordinal state transition models, proportional-odds models at a fixed time point, days-to-recovery scale analyses, Cox models for time-to-event endpoints, logistic regression for binary endpoints, generalized pairwise comparisons for hierarchical composites, and t-tests for days alive and out of hospital. This study will provide a systematic comparison of endpoint definitions and analysis methods for ARVI trials under clinically motivated data-generating mechanisms. The results are intended to inform the selection of feasible, interpretable, and statistically efficient primary analysis strategies for future trials in viral respiratory disease.

## Background

Acute viral respiratory infections (ARVIs) are among the most prevalent diseases and are a leading cause of hospitalizations and mortality worldwide [1, 2]. ARVIs are caused by a diverse set of viruses; among these, respiratory syncytial virus (RSV), influenza, and SARS-CoV-2 are pathogens of particular concern [3–5].

Substantial efforts have been made to identify effective treatments for hospitalized participants with ARVIs with limited success [5]. An important challenge when designing a randomized controlled clinical trial (RCT) in this participant population is deciding which outcomes to collect and how to analyze these to determine whether a treatment is effective. In particular, clinical trajectories of participants with ARVIs are not captured by a binary outcome such as alive versus dead: intermediate clinical states such as symptom burden, oxygen requirement, ventilation status or hospital discharge are often of direct relevance to participants [6]. Additionally, while mortality is an unambiguous and clinically important endpoint, it may be infeasible as a primary endpoint for clinical trials in an inter-pandemic setting due to low-event rates, which would require very large sample sizes for adequate statistical power [7, 8]. Reflecting this challenge, the European Medicines Agency recently held a virtual workshop to discuss feasible and clinically relevant primary efficacy endpoints for ARVIs [9].

Several endpoint strategies that incorporate information beyond death have been used or proposed in ARVI trials [6, 10]. For example, the Days to Recovery Scale at 60 days (DRS-60), developed for the STRIVE study, combines time-to-recovery with non-recovery states and death by mapping outcomes to an ordinal scale [10]. DRS-60 is a version of the STRIVE Clinical Recovery Scale that encodes time out of hospital as an ordinal variable time while penalizing rehospitalizations and death: values of 0 indicate the best outcomes, 60 indicates recovery on day 60, participants not recovered by day 60 are coded as 61, and death is coded as 62 [10]. A related strategy is to analyze time to sustained recovery, which has previously been defined as at least three consecutive days without symptoms and has been used as the primary outcome in multiple RCTs [11–13]. Ordinal severity scales evaluated at fixed time points form another common family of endpoints, in particular variants of the WHO clinical progression scale [14, 15].

More recently, Markov ordinal state transition models (MOST) have been proposed to analyse ordinal clinical states, such as the WHO clinical progression scale, longitudinally rather than restricting the analysis to a single assessment time point [16]. Recently, REMAP-CAP adopted MOST as the primary analysis method [17, 18]. REMAP-CAP uses a Bayesian approach; to date, MOST have not been used in a clinical trial of ARVIs with a frequentist framework. MOST is also currently considered in the SYNDACT trial [19, 20] as primary analysis.

Dodd et al. [6] investigated power for time-to-event outcomes and for ordinal severity scales evaluated at fixed time points. They found that power for fixed time-point methods depended strongly on the selected analysis time, whereas time-to-event approaches achieved reasonable power even when compared with a fixed-time-point analysis at its maximum. These results motivate a more systematic comparison of endpoint definitions and analysis strategies under data-generating mechanisms that reflect plausible participant trajectories.

Clinical trial simulations generate artificial data of clinical studies in computer programs that involve random number sampling and assumptions about an underlying data generating mechanism [21–24]. They are a flexible tool to investigate and compare statistical analysis approaches and are particularly valuable if analytical characterisations of the measure to be compared is difficult to find or if deviations from the typically assumed model is of interest.

Simulation protocols mitigate selective reporting of results and require the authors to make decisions regarding the design of the simulation study explicit and to justify these decisions [21]. Recently, this practice is adopted and several protocols for clinical trial simulation studies were published [25–27].

The aim of this study protocol is to detail a simulation study to investigate the type I error and power of commonly used and recently proposed approaches for ARVIs endpoints under a range of data-generating mechanisms designed to mimic clinically realistic trajectories. This protocol describes a simulation study which that has not yet generated operating-characteristic results. Participant recruitment is not applicable, because no prospective participants will be enrolled. These simulations are expected to be completed by October 2026. Results are expected to be available by December 2026.

In Section Simulation Design, the design of the simulation study is detailed. An overview of the planned scenarios is given in Section Overview of scenarios. Some technical considerations for the simulations are discussed in Section Technical considerations followed by consideration of limitations, reporting, and final remarks (Section Final Remarks).

## Simulation Design

Following the guidelines of Morris et al. [21], we describe five key components of our simulation study: Aims, Data-generating mechanisms, Targets (Estimands), Methods, and Performance measures (ADEMP).

### Aims (A)

We aim to compare different analysis approaches -i.e., different endpoint definitions and suitable statistical analysis methods - for a parallel-group, two-arm RCT in populations with ARVIs, with respect to the probability of declaring success, that is, rejection of the null hypothesis *H*_0_ in favour of treatment benefit:

- under different data generating mechanisms,
- under different underlying treatment effect sizes,
- for different durations of follow-up,
- for different types of populations: severely ill and moderately ill participants,
- for different total sample sizes.

See Section Overview of scenarios for an overview of scenarios studied.

### Data Generating Mechanisms (D)

We will generate data with *K* ∈ {4, 6, 8} states. We simulate 8-level longitudinal ordinal outcomes as used in the Adaptive COVID-19 Treatment Trials 1 and 2 (ACTT-1, ACTT-2) [28, 29], see Table 2. We then collapse the 8-level ordinal variable to obtain corresponding 6-level outcomes, as will be used in the EU-Syndact trials [20], and 4-level outcomes as was used in [30]; see Table 3 and Table 4. We do not consider outcomes with more levels, such as the 10 level WHO scale [31], as additional levels are of unclear clinical relevance. For example, additional levels from the WHO scale include viral load measurements, which are not a patient-centered outcome and are usually not collected daily. Scale-specific state sets used to construct derived endpoints, including recovered status, discharge, days alive and out of hospital, ventilation, death, and composite failure, are defined in Supplementary Table S5.

**Table 1.** Abbreviations used in the manuscript.

| Abbreviation | Definition |
| --- | --- |
| ACTT-1 | Adaptive COVID-19 Treatment Trial 1 |
| ACTT-2 | Adaptive COVID-19 Treatment Trial 2 |
| ADEMP | Aims, Data-generating mechanisms, Estimands, Methods, and Performance measures |
| ARVI | Acute viral respiratory infection |
| CDF | Cumulative distribution function |
| COVID-19 | Coronavirus disease 2019 |
| DC | Design choice |
| DC(c) | Constrained design choice |
| DGM | Data-generating mechanism |
| DOH | Days alive and out of hospital |
| DRS-60 | Days to Recovery Scale at 60 days |
| DRS-H | Days-to-Recovery Scale through follow-up horizon $H$ |
| DSF | Disease-specific feature |
| DSF(a) | Assumed disease-specific feature |
| DSF(e) | Estimable disease-specific feature |
| ECMO | Extracorporeal membrane oxygenation |
| EMA | European Medicines Agency |
| EU | European Union |
| GPC | Generalized pairwise comparisons |
| $H_0$ | Null hypothesis |
| $H_A$ | Alternative hypothesis |
| HR | Hazard ratio |
| MC-CI | Monte Carlo confidence interval |
| MCSE | Monte Carlo standard error |
| MOST | Markov ordinal state transition model |
| NTB | Net treatment benefit |
| NIH | National Institutes of Health |
| NIV | Non-invasive ventilation |
| NTB | Net treatment benefit |
| OR | Odds ratio |
| PO-H | Proportional-odds analysis at day $H$ |
| RCT | Randomized controlled trial |
| RSV | Respiratory syncytial virus |
| SARS-CoV-2 | Severe acute respiratory syndrome coronavirus 2 |
| SE | Standard error |
| US | United States |
| VGAM | Vector generalized linear and additive models; R package name used for ordinal models |
| WHO | World Health Organization |

**Table 2.**
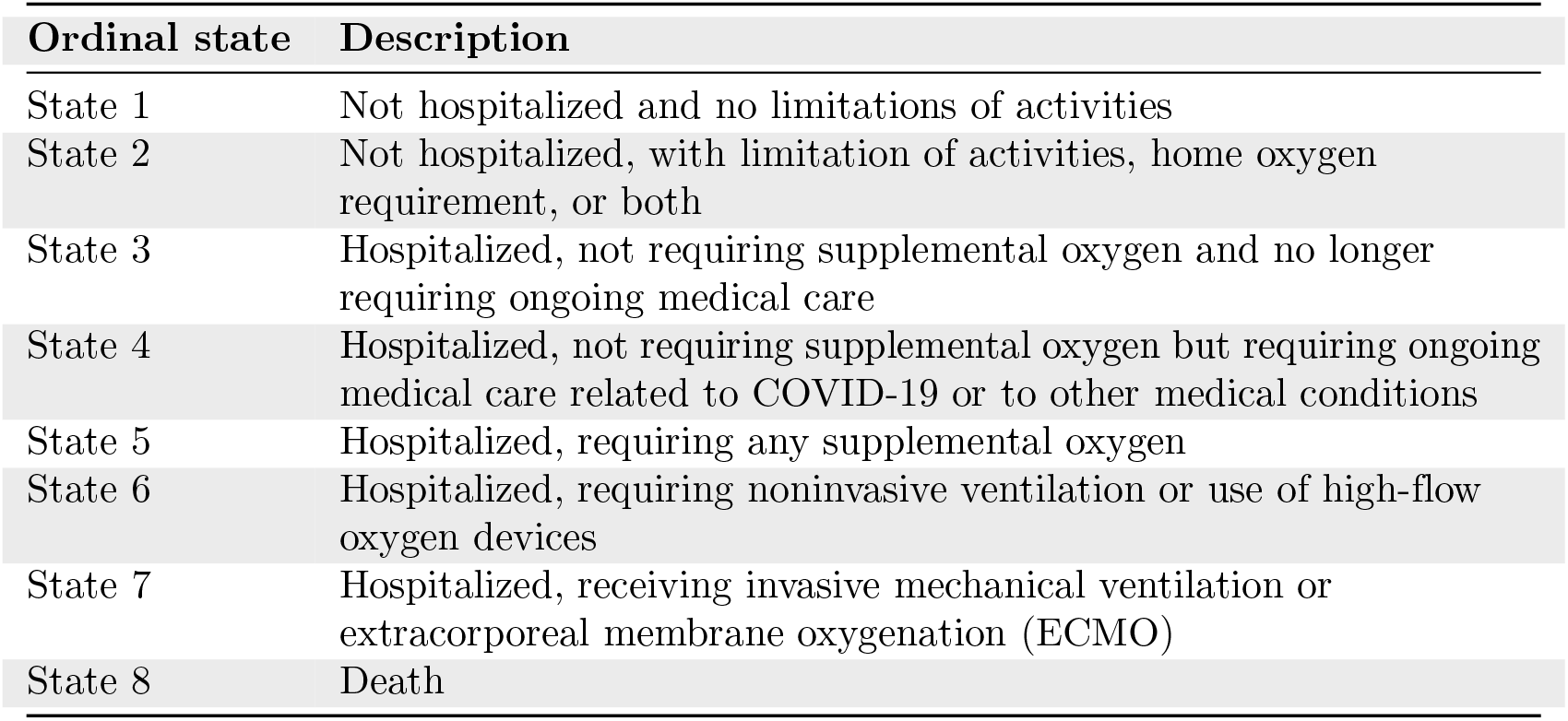
Ordinal outcome scale used in ACTT-1/ACTT-2.

**Table 3.** Ordinal outcome scale used in EU-Syndact and mapping from ACTT categories.

| Ordinal state | Description | ACTT |
| --- | --- | --- |
| State 1 | Not hospitalized | 1–2 |
| State 2 | Hospitalized, no oxygen therapy | 3–4 |
| State 3 | Hospitalized, minimal organ support (e.g., nasal oxygen) | 5 |
| State 4 | Hospitalized, intermediate organ support (e.g., NIV, High-flow) | 6 |
| State 5 | Hospitalized, intensive organ support (e.g., Intubation, ECMO, Dialysis) | 7 |
| State 6 | Death | 8 |

**Table 4.** VIOLET ordinal outcome scale and mapping from ACTT categories.

| Ordinal state | Description | ACTT |
| --- | --- | --- |
| State 1 | Not hospitalized | 1–2 |
| State 2 | Hospitalized, not treated for acute respiratory distress syndrome | 3–5 |
| State 3 | Hospitalized, treated for acute respiratory distress syndrome | 6–7 |
| State 4 | Dead | 8 |

Let *Y*_*it*_ ∈ {1, 2, …, *K*} denote the daily state generated on the ACTT scale for participant *i* = 1, …, *N*, with *N* being the total number of participants in the RCT, at time *t* = 0, …, *H*, where *t* are days since randomization with *t* = 0 corresponding to the day of randomization and *H* is the maximum duration of follow-up common to all participants.

For all mechanisms, *i* = 1, …, *N* indexes participants, *t* = 0, …, *H* indexes time, and *Y*_*it*_ ∈ *S* = {1, …, *K*} is the ordinal state (1 = best, *K* = worst). Let *A*⊊*S* denote the set of absorbing states (e.g., *A*= {*K*}, death). Once a participant enters a state in *A*, they remain in that state for all subsequent time points. For all data generating mechanisms, treatment is defined as *Z*_*i*_ ∈ {0, 1}, where *Z*_*i*_ = 1 indicates being part of the active treatment arm. The parameters required to implement each data-generating mechanism, scenario-specific treatment-effect parameters, absorbing states, and event-to-state mappings, are summarized in Supplementary Table S1.

A superpopulation of at least 250,000 participants is generated and the true treatment effect determined by calculating the between-group difference for the estimand of interest for each method. The difference in time alive and out of hospital between the groups defines the treatment effect for each scenario. For each analysis method, the corresponding method-specific estimand will also be estimated in the large superpopulation and reported to aid interpretation; however, these method-specific estimands will not be used to define the scenario labels. See also Section Overview of scenarios. In each simulation repetition, *N/*2 participants are sampled to each group.

To support the reader with notation, symbols used and their definition are listed in Table 5.

**Table 5.** Table of notation and symbols.

| Notation or symbol | Description |
| --- | --- |
| $K$ | Number of states in a scale definition |
| $k$ | Category or state index |
| $\mathcal{S}$ | State space, ($\mathcal{S} = \{1, \dots, K\}$) |
| $\mathcal{A}$ | Set of absorbing states in the scale K. Usually the highest state K (death) |
| $i$ | Participant index |
| $t$ | Time index (day) |
| $H$ | Observation/Follow-up period |
| $n_T, n_C, N$ | Treatment-group sample size, control-group sample size, and total sample size in one simulated trial |
| $Y_{it}$ | State of participant $i$ at day $t$ |
| $Z_i, z$ | Treatment indicator (of participant $i$) |
| $L_{it}$ | Latent variable in the Brownian motion DGM |
| $\sigma_{L0}$ | Standard deviation of baseline latent severity in the Brownian motion DGM |
| $b_i$ | Participant-specific random drift effect in the latent Brownian motion DGM |
| $\sigma_b$ | Standard deviation of the participant-specific random drift effect in the latent Brownian motion DGM |
| $\mu_i(t), w(t), \tilde{\mu}_i(t)$ | Participant- and time-dependent drift, attenuation function, and effective drift in the latent Brownian-motion DGM |
| $\mu_0, \mu_1, \mu_2, \mu_Z$ | Fixed effect drift parameters in the latent Brownian-motion DGM |
| $c_1, c_2$ | Time points at which drift attenuation begins and reaches zero |
| $\varepsilon_{it}$ | Innovation term in the latent Brownian-motion DGM |
| $\sigma_{rw}$ | Standard deviation of the innovation term in the latent Brownian-motion DGM |
| $\delta_k, \gamma_k$ | Thresholds/cutpoints in the latent Brownian-motion DGM and ordinal Markov DGM, respectively |
| $F(u)$ | Logistic cumulative distribution function |
| $\rho$ | Potential transition rate parameter in the latent Brownian-motion DGM |
| $r$ | Index of potential transition intervals |
| $G_{ir}$ | Waiting time until the $r$-th potential transition for participant $i$ |
| $R_{ir}$ | $r$-th potential transition day for participant $i$ |
| $\mathcal{R}_i$ | Set of potential transition days for participant $i$ |
| $N(0, \sigma^2)$ | Normal distribution with mean 0 and variance $\sigma^2$ |
| $e$ | Endpoint index |
| $\boldsymbol{\beta}, \beta$ | (Vector of) model coefficients (subscripts indicate associated variables) |
| $\mathbf{x}_i$ | Vector of covariates of participant $i$ |
| $m, M$ | Occurrence of an event and maximum number of events for a latent piecewise exponential (LPE) recurrent event |
| $W_k^{(i)}$ | Waiting time in between events of type $k$ for participant $i$ in the LPE DGM |
| $Exp(\lambda)$ | Exponential distribution with parameter $\lambda$. |
| $E_{k,m}^{(i)}, \mathcal{E}_i$ | Event time of event type $k$ (state $k$), event occurrence $m$ and participant $i$ and the ordered set of those times, respectively, in the LPE DGM |
| $h(t)$ | Hazard function |
| $HR$ | Hazard ratio |
| $H_0, H_A$ | Null and alternative hypothesis |
| $\alpha$ | Nominal significance level |
| $\mathbb{1}(\dots)$ | Indicator function for condition $(\dots)$ |
| $\mathcal{R}$ | Set of recovered states in DRS score |
| $R_i$ | Condition to declare a trial a success |
| $n_{\text{sim}}$ | Number of simulation repetitions |
| $\alpha_k$ | Cumulative threshold of category/state $k$, state-specific intercept of the linear term |
| $\mathcal{B}$ | Set of states that define the respective binary endpoint |
| $DOH_{i,H}, \overline{DOH}_Z$ | Days alive and out of hospital for participant $i$ and follow-up $H$ and respective group mean in treatment arm $z$ |
| $wins, losses, ties$ | Number of wins, losses and ties, and number of all possible pairs |
| $N_{\text{pairs}}$ | from GPC |
| $NTB$ | Net treatment benefit |
| $p_r, \hat{p}_r$ | Rejection probability and according estimate, respectively |
| $p_{\text{fail}}, \hat{p}_{\text{fail}}$ | Convergence failure rate and according estimate |
| $s$ | Simulation repetition |
| $f_s$ | indicator of convergence failure in repetition $s$ |
| $MCSE$ | Monte Carlo standard error |
| $C_i$ | Indicator of successful model convergence |

#### Mechanism 1: Latent Brownian Motion

This mechanism uses a latent severity process *L*_*it*_ on a logistic scale, with *i* and *t* as defined above. The latent severity determines daily state probabilities through a latent-threshold model, but observed non-absorbing state changes are only allowed (but not forced) on random transition time points. Although the latent process itself is a first-order Markov process, many latent values map to the same observed state.

Therefore, conditioning on *Y*_*i,t*™1_ does not fully recover *L*_*i,t*™1_, and thus violates the first-order Markov assumption underlying MOST. Data generation with a latent Brownian motion process is therefore suitable to evaluate the Type I error rate of MOST when the first-order Markov condition does not hold. Because this mechanism also allows us to simulate plausible participant trajectories, it serves as the primary data generating mechanism. In the following, we describe this data generating mechanism in detail.

Baseline latent severity is 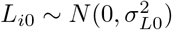 . We include a drift term so that latent health can systematically improve or worsen over time, and also level off later in follow-up. The drift terms may be common to all cumulative thresholds or specified separately for each of the *K* ™ 1 thresholds. To allow for participant-specific latent drift, we introduce a random drift 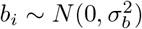, independent across participants. 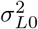 and 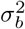 are the variances specific to the respective processes. For *t* ≥ 1, we define the unattenuated drift

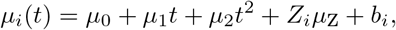

Systematic drifts such as disease progression may not be constant over time. For instance, a participant’s health might change more in the initial phase of a study, or might naturally stabilize after a period of decline. To capture this, we introduce a time-dependent attenuation function, *w*(*t*) which modulates the strength of the drift term over the course of the follow-up period:

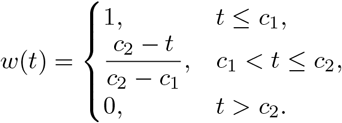

The effective drift applied at time *t* is 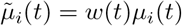 . Here, before *c*_1_ the drift has full effect and then declines gradually until after *c*_2_ the drift is zero.

To represent day-to-day fluctuation around this average drift, we add random fluctuations 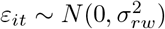 which are independent across participants and time points. When drift parameters are common across thresholds, the latent severity for *t* ≥ 1 is 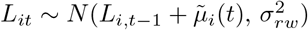 . When threshold-specific parameters are used, the update is applied using the average drift across thresholds.

We use baseline thresholds *δ*_*k*_ on the latent logistic scale to sample the observed state *Y*_*it*_, where *δ*_1_ *> δ*_2_ *>*· · · *> δ*_*K*™1_. Let *F* (*u*) = {1 + exp(™ *u*)} ^™1^ be the logistic cumulative distribution function (CDF). When the drift parameters are common across thresholds, these thresholds remain fixed, and time and treatment act thought the latent severity process *L*_*it*_. When threshold-specified drift parameters are used, the same latent severity process is applied, but the thresholds used in the cumulative logits are allowed to vary over time and by treatment arm. Specifically, each baseline threshold is shifted according to the accumulated drift for that threshold, relative to the average drift used to update the latent severity.

The cumulative probability of being in state *k* or higher is thus

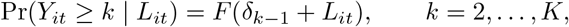

In the common-drift case, *δ*_*k*™1_ is the fixed baseline threshold. In the threshold-specific case, *δ*_*k*™1_ denotes the threshold value used for participant *i* on day *t* in treatment arm *Z*_*i*_, after applying the accumulative threshold-specific time and treatment effects.

Because state 1 is best and state *K* is worst, a larger latent severity *L*_*it*_ increases Pr(*Y*_*it*_ ≥ *k* | *L*_*it*_) and shifts probability mass to worse states.

State probabilities are obtained by differencing:

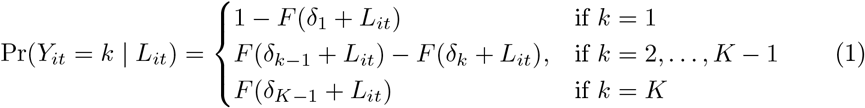

In the threshold-specific case, each threshold term is evaluated after applying the accumulated time and treatment effects for that threshold. At baseline, the observed state *Y*_*i*0_ is sampled from these cell probabilities evaluated at *L*_*i*0_, optionally restricted to a prespecified admissible baseline state set (e.g., *S\A*) by renormalizing probabilities on that set.

To reduce the number of observed non-absorbing state transitions, each participant is assigned random transition times. Let

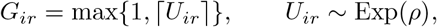

where *ρ* > 0 is the potential transition rate parameter and *r* = 1, 2, … indexes potential transition intervals. The transition days are

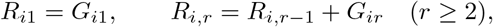

and we write *R*_*i*_ = {*R*_*i*1_, *R*_*i*2_, … } for the set of potential transition days for participant

Generation proceeds as follows:

In this DGM, the only absorbing state is death, so *A* = {*K*}. At *t* = 0, sample *L*_*i*0_ and then sample *Y*_*i*0_ from the baseline state probabilities. For *t* ≥ 1, if *Y*_*i,t*™1_ ∈ *A*, set *Y*_*it*_ = *Y*_*i,t*−1_ and do not update the latent process further.

Otherwise, first update the latent severity:

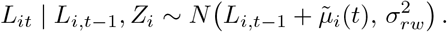

Entry into the absorbing state is allowed on any day, not only on potential transition days. Thus, if *K* is absorbing (e.g., death), the participant enters state *K* on day *t* with probability Pr(*Y*_*it*_ = *K* | *L*_*it*_).

If death does not occur on day *t* and *t*∈*R* _*i*_, a new non-absorbing state is sampled from the conditional distribution

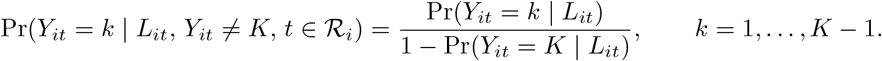

If death does not occur and *t* ∉ *R*_*i*_, the previous observed state is carried forward, that is, *Y*_*it*_ = *Y*_*i,t*−1_.

This construction yields a latent process that evolves daily, while the observed non-absorbing state changes only intermittently. The resulting trajectories are expected to exhibit fewer short-term state changes, especially fewer home-to-hospital transitions after discharge, while still allowing daily deterioration to death.

#### Mechanism 2: Ordinal Markov Process

This mechanism generates trajectories using a discrete-time first-order Markov chain where transition probabilities depend on the previous state, treatment assignment, baseline covariates, and time, under a proportional odds model. This mechanism allows us to simulate realistic participant state data because the parameter estimates used can be obtained from historical data. However, under this mechanism, the first-order Markov assumption is true per definition, and is expected to favour MOST.

Let ***x***_*i*_ denote a vector of baseline covariates (e.g., age, disease severity scores), sampled from empirically informed distributions. Baseline states *Y*_*i*0_ are sampled together with other baseline covariates from a baseline distribution reflecting typical admission states.

Let *F* (*u*) be the logistic CDF as above.

For *t* ≥ 1, define the linear predictor

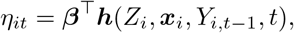

where ***h***(·) is a function encoding treatment, covariates, previous state indicators, time, and potentially interactions (e.g., previous state *×* time). The proportional odds model gives cumulative probabilities

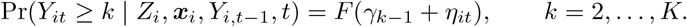

We define *γ*_1_ > *γ*_2_ > > *γ*_*K*−1_, so that cumulative probabilities are non-increasing in *k* and resulting cell probabilities are nonnegative. Cell probabilities are obtained by differencing:

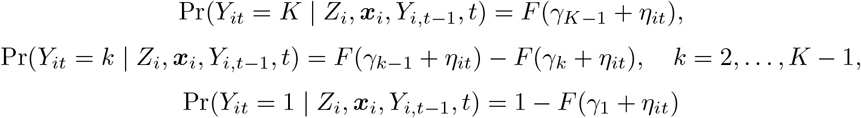

Generation proceeds as follows: for *t* = 0 (baseline), sample *Y*_*i*0_ from the baseline distribution.

For *t* ≥ 1:

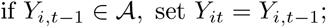

otherwise, sample *Y*_*it*_ from the cell probabilities above conditional on *Y*_*i,t*−1_ and ***x***_*i*_.

#### Mechanism 3: Latent piecewise exponential event times

The third mechanism is based on treating state transitions as independent recurrent events with exponentially distributed waiting times between them. The cumulative sum defines the times at which the participant enters the respective state. If an absorbing state is reached, the participant stays in this state until the end of follow-up. By introducing participant-level frailty terms, the course of a participant is correlated for the whole observation period and breaks the Markov assumption. Further, for some applications, considering the participant state as event times is natural, e.g., time of death, or time of deterioration of lung function and start of mechanical ventilation.

For simplicity in coding, a maximum number, *M*, of events is set and all these event times generated.

Let

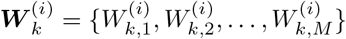

be a vector of waiting times for the participant *i* and the event type *k*, with 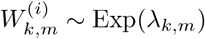, where Exp(*λ*) is the exponential distribution and parameters *λ*_*k,m*_ may differ depending on the event type and the number of previous events. Let

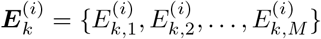

with 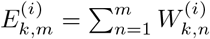 denote the event times for the participant *i* and event type *k*. All event times for participant *i* are then given by the ordered set 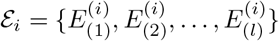, obtained by listing event times across event types and ignoring event times after an absorbing event occurs. Thus, 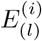 either denotes the last event time within follow-up or the time at which the participant enters an absorbing state. The states for participant *i* at time *t* are then defined by the event type with the most recent event time and the state only changes if a new event type occurs.

The floor function is applied to continuous event times to get the day of the event. Continuous event times are ordered before applying the floor function. If two or more events occur on the same day, only the last event is recorded, unless one of the events is death which is always recorded and absorbing. If an absorbing state is reached within day zero (recruitment day), the event is only recorded the next day (day 1).

The treatment effect is implemented by multiplying the rates *λ*_*k,m*_ of waiting-time distributions of participants in the treatment group by event-type-specific hazard ratios *HR*_*k*_.

To introduce dependence between event types, a participant-level frailty term is added that influences the event-time generation of all or some event types of a participant. This frailty term is assumed to follow a normal distribution with expectation 0 and is treated like a covariate that has different influence on events of different event types. Let *u*_*i*_ ∼ *N* (0, *σ*) be the participant-level frailty. Then the hazard for participant *i*, and event occurrence *m* of event type *k, λ*_*i,k,m*_, is

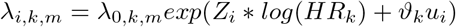

where the magnitude of ϑ_*k*_ determines the influence of the frailty term and the sign determines the direction of the dependence between frailty and event type. Negative values are set for favourable event types (like discharge) and positive values for unfavourable event types (like death). In this way, frail participants tend to experience unfavourable outcomes more often.

#### Resampling from ACTT-2 data

To investigate empirical operating characteristics on real data we use individual participant data from the ACTT-2 trial consisting of 1033 participants, which collected daily participant-state data. ACTT-2 investigated baricitinib plus remdesivir in hospitalized adults with COVID-19 [29]. We treat the control (518 participants) and intervention group (515 participants) as two distinct empirical populations.

For each simulation iteration treatment and control group are of equal size (*n*_*T*_ [treatment] = *n*_*C*_ [control]). Participants are then selected by randomly sampling with replacements from the empirical populations.

- Under *H*_0_: To simulate a scenario with no treatment effect, participants for both groups are sampled *with replacement* from the ACTT-2 control arm.
- Under *H*_*A*_: To evaluate the methods under the treatment effect observed in ACTT-2, i.e., to evaluate the empirical power, *n*_*T*_ participants are sampled *with replacement* from the original ACTT-2 treatment arm, and *n*_*C*_ participants are sampled *with replacement* from the original control arm.

Other parameters, such as follow-up time, state scale and treatment effects are detailed in Section Overview of scenarios.

### Estimands and other Targets (E)

The approaches compared in this simulation study are combinations of different endpoint definitions and suitable statistical analysis methods. In this way, they target different estimands and associated treatment effect estimates (e.g., marginal odds ratios at a fixed time point for dichotomous endpoints, hazard ratios for a time-to-event endpoints, mean difference in continuous endpoints such as days out of hospital). Thus, it will be difficult to directly compare the different approaches.

Instead, our primary focus is on the first of Senn’s five questions for clinical trials: *“Was there an effect of treatment in this trial?”* [32]. The target of the simulation study is the unconditional rejection rate of the analysis-approach-specific null hypothesis. Accordingly, for each approach and each follow-up horizon *H* ∈ {28, 60}, we define “success” as rejection of the respective test’s null hypothesis of no or unfavourable treatment effect of the treatment at level *α* = 0.025 (one-sided). For all methods, success will be defined as an estimated treatment effect in the prespecified favourable direction together with a one-sided p-value < 0.025. Equivalently, when only two-sided software p-values are available, success will require the estimate to be in the favourable direction and the two-sided p-value to be < 0.05.. Operating characteristics are summarized by the probability of declaring success (type I error under the *H*_0_ scenarios; power under the *H*_*A*_ scenarios) as discussed in Section Performance measures used for evaluating the method (P), Table 6 and Supplementary Table S4. If an analysis method fails to deliver a valid decision in a simulation iteration, the analysis method is considered to declare trial success if the target is Type-I-error or is considered as not declaring the trial a success if the target is statistical power (worst-case imputation of missing decisions). A secondary target is conditional on the respective analysis method providing a valid trial decision of the iteration. The number of failed iterations for each scenario and analysis approaches will be reported. See also Section Technical considerations on handling of convergence and other failures.

**Table 6.**
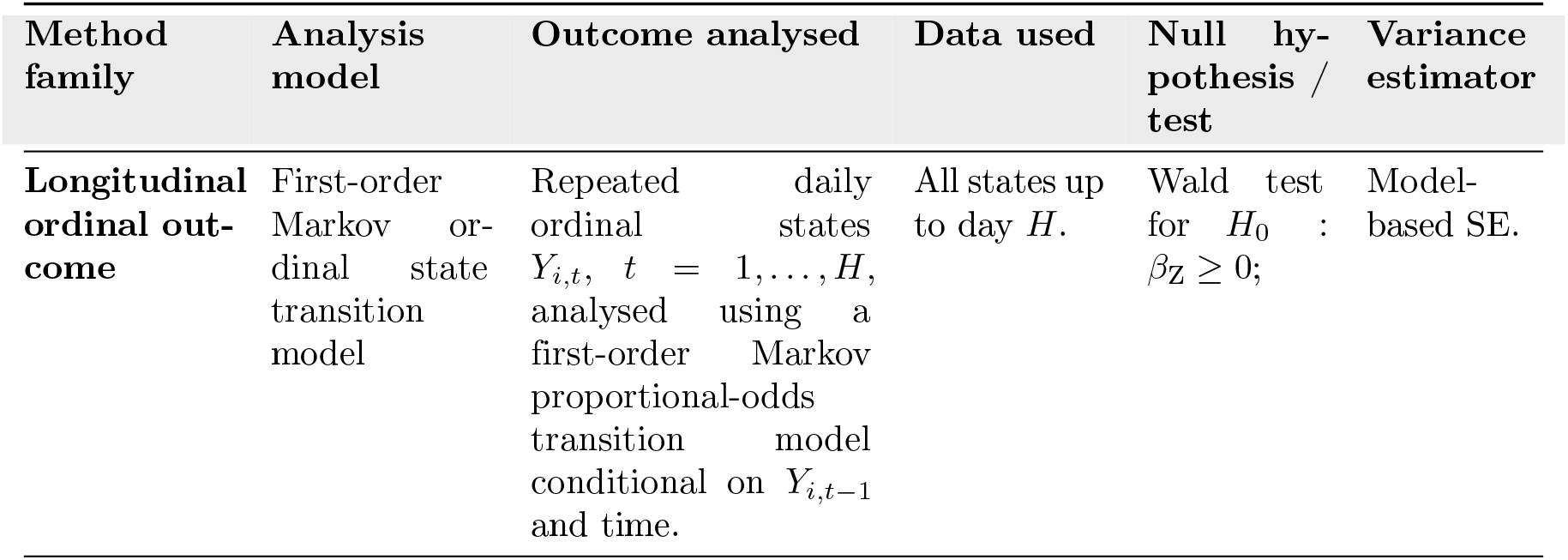
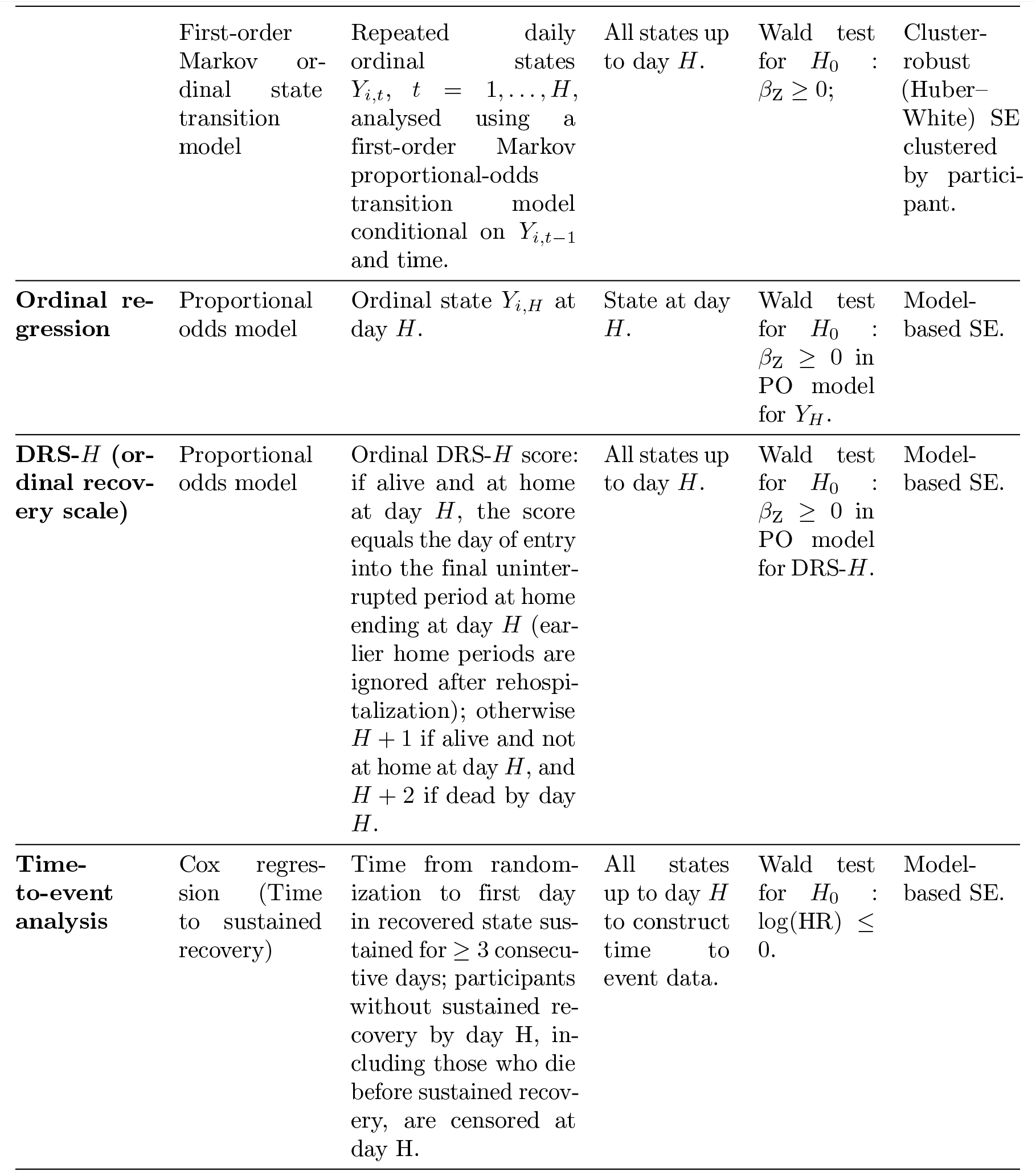
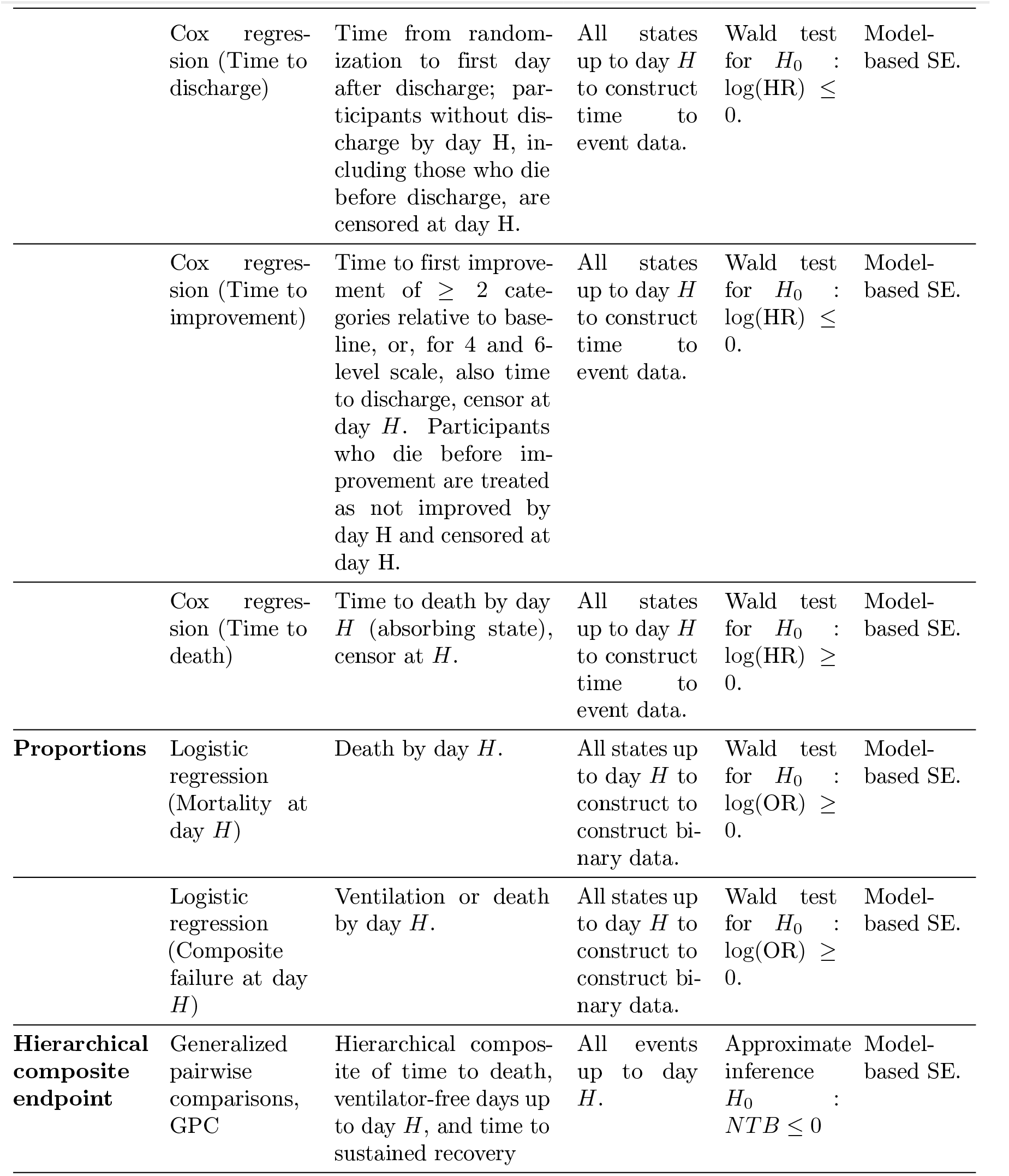
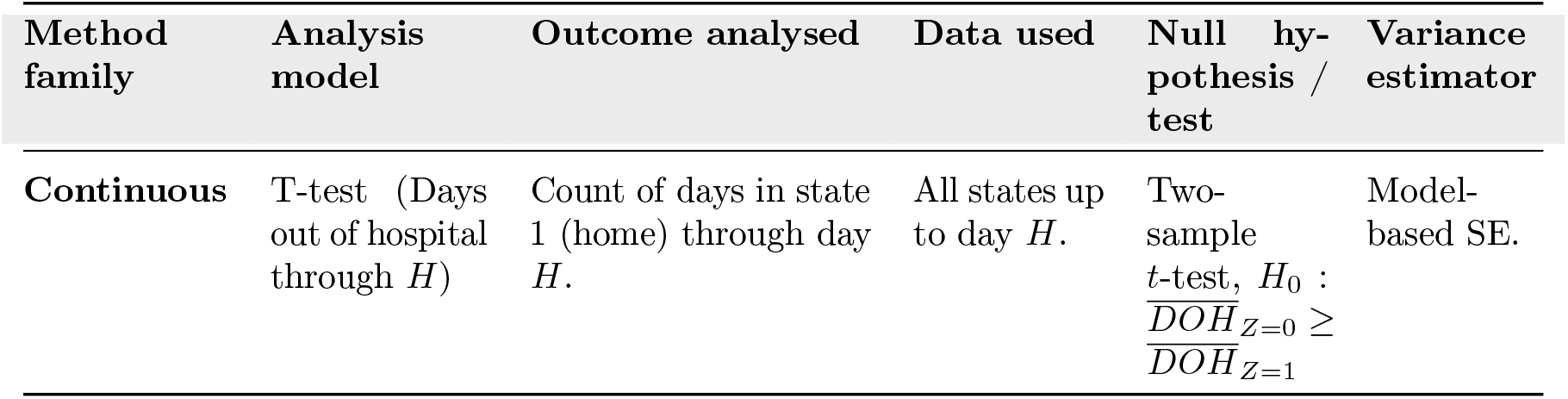
Analysis methods, analyzed outcomes, null hypotheses, and variance estimators evaluated at horizons *H* ∈ {28, 60} . Null hypotheses assume that *Z*_*i*_ = 1 in the treatment group. 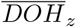 is the group mean of time alive and out of hospital in treatment arm *z*.

| Method family | Analysis model | Outcome analysed | Data used | Null hypothesis / test | Variance estimator |
| --- | --- | --- | --- | --- | --- |
| Longitudinal ordinal outcome | First-order Markov ordinal state transition model | Repeated daily ordinal states $Y_{i,t}$, $t = 1, \dots, H$, analysed using a first-order Markov proportional-odds transition model conditional on $Y_{i,t-1}$ and time. | All states up to day $H$. | Wald test for $H_0 : \beta_Z \geq 0$; | Model-based SE. |
| | First-order Markov ordinal state transition model | Repeated daily ordinal states $Y_{i,t}$, $t = 1, \dots, H$, analysed using a first-order Markov proportional-odds transition model conditional on $Y_{i,t-1}$ and time. | All states up to day $H$. | Wald test for $H_0 : \beta_Z \geq 0$; | Cluster-robust (Huber–White) SE clustered by participant. |
| **Ordinal regression** | Proportional odds model | Ordinal state $Y_{i,H}$ at day $H$. | State at day $H$. | Wald test for $H_0 : \beta_Z \geq 0$ in PO model for $Y_H$. | Model-based SE. |
| **DRS-$H$ (ordinal recovery scale)** | Proportional odds model | Ordinal DRS-$H$ score: if alive and at home at day $H$, the score equals the day of entry into the final uninterrupted period at home ending at day $H$ (earlier home periods are ignored after rehospitalization); otherwise $H + 1$ if alive and not at home at day $H$, and $H + 2$ if dead by day $H$. | All states up to day $H$. | Wald test for $H_0 : \beta_Z \geq 0$ in PO model for DRS-$H$. | Model-based SE. |
| **Time-to-event analysis** | Cox regression (Time to sustained recovery) | Time from randomization to first day in recovered state sustained for $\geq 3$ consecutive days; participants without sustained recovery by day $H$, including those who die before sustained recovery, are censored at day $H$. | All states up to day $H$ to construct time to event data. | Wald test for $H_0 : \log(\text{HR}) \leq 0$. | Model-based SE. |
| | Cox regression (Time to discharge) | Time from randomization to first day after discharge; participants without discharge by day $H$, including those who die before discharge, are censored at day $H$. | All states up to day $H$ to construct time to event data. | Wald test for $H_0 : \log(\text{HR}) \leq 0$. | Model-based SE. |
| | Cox regression (Time to improvement) | Time to first improvement of $\geq 2$ categories relative to baseline, or, for 4 and 6-level scale, also time to discharge, censor at day $H$. Participants who die before improvement are treated as not improved by day $H$ and censored at day $H$. | All states up to day $H$ to construct time to event data. | Wald test for $H_0 : \log(\text{HR}) \leq 0$. | Model-based SE. |
| | Cox regression (Time to death) | Time to death by day $H$ (absorbing state), censor at $H$. | All states up to day $H$ to construct time to event data. | Wald test for $H_0 : \log(\text{HR}) \geq 0$. | Model-based SE. |
| **Proportions** | Logistic regression (Mortality at day $H$) | Death by day $H$. | All states up to day $H$ to construct to construct binary data. | Wald test for $H_0 : \log(\text{OR}) \geq 0$. | Model-based SE. |
| | Logistic regression (Composite failure at day $H$) | Ventilation or death by day $H$. | All states up to day $H$ to construct to construct binary data. | Wald test for $H_0 : \log(\text{OR}) \geq 0$. | Model-based SE. |
| **Hierarchical composite endpoint** | Generalized pairwise comparisons, GPC | Hierarchical composite of time to death, ventilator-free days up to day $H$, and time to sustained recovery | All events up to day $H$. | Approximate inference $H_0 : NTB \leq 0$ | Model-based SE. |
| **Continuous** | T-test (Days out of hospital through $H$) | Count of days in state 1 (home) through day $H$. | All states up to day $H$. | Two-sample $t$-test, $H_0 : \overline{DOH}_{Z=0} \geq \overline{DOH}_{Z=1}$ | Model-based SE. |

### Methods to be compared (M)

Table 6 summarizes all analysis methods. For each method, analyses are performed at prespecified analysis horizons, and treatment effects are evaluated using the method-specific favourable direction and one-sided *α* = 0.025 decision rule summarized in Table 6 and specified in detail together with method-specific estimands, favourable directions, null hypotheses, variance estimators, and implementation rules Supplementary Table S4.

### Markov ordinal state transition model (MOST)

MOST has previously been described in detail [16]. Compared with single-time-point proportional-odds models or standard Cox models, MOST requires more explicit modeling choices, including the functional form of time, how state history enters the model, and how inference is handled under repeated within-participant observations. For this simulation, we use a specification close to Rohde et al. [16], simplified to represent a pragmatic baseline analysis for ARVI trials.

Specifically, we fit a first-order Markov proportional-odds model to all observed daily states up to post-randomization day *H*, conditioning on previous-day state, treatment assignment, and follow-up time.

For participant *i* at day *t*, conditional on the previous day’s state *Y*_*i,t*−1_, the model has the ordinal cumulative logit form

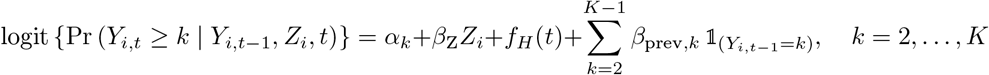

Model fitting is restricted to days with *Y*_*i,t*−1_ ∉ *A*_*K*_, since outcomes after entry into an absorbing state are deterministic.

where

- *α*_*k*_ indexes cumulative threshold *k* in the analysis model,
- *β*_Z_ represents the treatment effect,
- *f*_*H*_ (*t*) is a restricted cubic spline in follow-up time, with the spline basis depending on the analysis horizon *H*,
- *β*_prev,*k*_ represents the effect of previous-day state *k* relative to the reference state 1; because model fitting excludes absorbing previous states, this yields *K* − 2 previous-state coefficients when state *K* is absorbing.

For analyses up to day 28, *f*_*H*_ (*t*) uses 5 knots at empirical quantiles (0.05, 0.275, 0.50, 0.725, 0.95). For analyses up to day 60, *f*_*H*_ (*t*) uses 6 knots at empirical quantiles (0.05, 0.23, 0.41, 0.59, 0.77, 0.95). We do not relax the proportional odds assumption for time. When we applied MOST to real-world data from two RCTs in COVID-19 [29, 33], relaxing the proportional odds assumption for time only resulted no or only marginally better fit and had virtually no impact on model derived occupancy probabilities.

The treatment coefficient *β*_Z_ has a conditional interpretation: it is the log-odds ratio for being in a worse state on day *t*, conditional on the previous day’s state and the modelled time effect. It is therefore not directly comparable to marginal treatment effects from methods that target a single-time-point or population-averaged estimand.

For a first-order transition model, dependence beyond the previous outcome is excluded. In RCTs, this is an unavoidable assumption because state history before baseline is usually unavailable.

Under this working model, model-based standard errors rely on correct specification of the transition model and induced dependence structure. In practice, residual within-participant dependence may remain even after conditioning on *Y*_*i,t*−1_ and *t*, which can make model-based standard errors anti-conservative. We therefore report both (i) model-based standard errors and (ii) participant-level cluster-robust (Huber–White) standard errors [34, 35]. In both variants, the null hypothesis is *H*_0_ : *β*_Z_ ≥ 0.

#### Proportional odds at day H (PO-H)

PO-*H* uses only the ordinal state at day *H*, denoted *Y*_*i,h*_. The model is

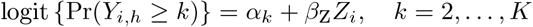

The treatment effect is summarized by a marginal proportional-odds log-odds ratio, and we test *H*_0_ : *β*_Z_ ≥ 0.

#### DRS-H (ordinal recovery scale)

For each participant *i*, daily states up to day *H* are mapped to an ordinal DRS-*H* score. Let *R* denote the set of recovered states for the scale being analysed. For the 4- and 6-level scales, recovered status for DRS-H is defined as being in state 1, *R* = {1} . For the 8-level ACTT scale, recovered status is defined as being in state 1 or state 2, *R* = {1, 2}. If a participant is alive and in a recovered state on day *H*, we define

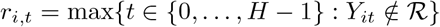

with *r*_*i,t*_ = 0 if no such day exists. The recovery day is then

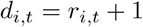

which is the start day of the final uninterrupted home period before day *H*.

The score is then

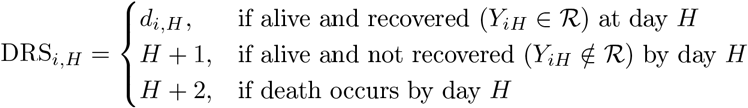

This definition implies that rehospitalization resets the counter: earlier home periods do not contribute once a later hospitalization occurs. Lower values indicate better outcomes. DRS-*H* is analyzed with a proportional-odds model with treatment as the only covariate, and the null hypothesis is *H*_0_ : *β*_Z_ ≥ 0.

#### Time-to-event endpoints

For each time-to-event endpoint, let 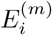 denote event time for endpoint *e* and *Z*_*i*_ = 1 indicates treatment group assignment. Analyses use Cox proportional hazards models

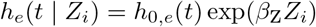

with Wald tests of *H*_0_ : *β*_Z_ ≥ 0 for unfavourable outcomes like death. For favourable time-to-event outcomes such as sustained recovery, discharge, and improvement, *H*_0_ : *β*_Z_ ≤ 0.

The endpoint definitions are: Favourable events: participants who do not experience the event by day *H* are censored at day *H*; participants who die before the event are treated as not having experienced the favourable event by day *H* and are censored at day *H*.

- **Time to sustained recovery:** first day in state 1 (4 and 6-level scale) and state 1 or 2 (8-level scale) followed by at least 3 consecutive days in state 1 or 1 and 2, respectively;
- **Time to discharge:** first day in state 1 (4 and 6-level scale) or one of states 1 or 2 (8-level scale);
- **Time to improvement:** first day with improvement of at least 2 ordinal categories relative to baseline state, or first day in state 1, if a 4 or 6-level scale is used and participants start in state 2

Unfavourable events:

- **Time to death:** first day in the absorbing death state; participants alive at day *H* are censored at day *H*.

#### Binary endpoints at day H

Two binary outcomes are evaluated at day *H*:

- **Mortality by day** *H*: indicator of death by day *H*.
- **Composite failure by day** *H*: indicator of invasive ventilation or death by day *H*.

Composite failure will be constructed from the underlying 8-level ACTT states before any scale collapse: failure is ACTT state 7 or 8 by day H, corresponding to invasive mechanical ventilation, ECMO or death. The same binary variable will be used for analyses using 6- or 4-level scales to avoid ambiguity introduced by scale collapse as the 4-state scale does not allow to distinguish invasive ventilation from other treatment of respiratory distress syndrome.

Both are analyzed with logistic regression

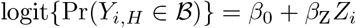

where *B* is the set of states that defines the binary endpoint, e.g., *B* = *A* for mortality by day *H*. The null hypothesis tested is *H*_0_ : *β*_Z_ ≥ 0 for *Z*_*i*_ = 1 in the treatment group.

#### Days out of hospital through day H

For each participant, time alive and out of hospital is defined as

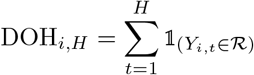

where *R* is the set of recovered states specific to the state scale as defined above.

Define Δ = *mean*(*DOH*_*i,H*_ |*Z*_*i*_ = 1) − *mean*(*DOH*_*i,H*_ |*Z*_*i*_ = 0). Group means are compared using a two-sample t-test, with directional null hypothesis *H*_0_ : Δ ≤ 0.

#### Hierarchical composite endpoints

A hierarchical composite endpoint ranks participants based on comparisons of outcomes in order of (clinical) priority and assessed until day *H*. Components of hierarchical endpoints in infectious disease trials commonly include mortality and further endpoints reflecting clinical failure (e.g., microbiological relapse, days spent in the intensive care unit), complications or adverse events (e.g.,secondary infections, mechanical ventilation), and recovery (e.g.,ventilator-free days, length of hospital stay, symptom resolution) [36–38]. Given that all participants in the current setting start in hospital, we define a hierarchical composite endpoint as

1. Time to death by any cause
  - Longer times are favourable
2. Days ventilator free and alive
3. Time to sustained recovery
  - Discharge from hospital
  - No re-hospitalisation in the following 3 days
  - Shorter times are favourable

in order of clinical priority. Further endpoint definitions may be explored in the simulations (e.g., including days alive and hospitalized).

The time-to-event endpoints will be censored at the end of follow-up, *H*, if no event occurred.

Hypothesis tests will be performed using generalized pairwise comparisons (GPC) [39]. In every possible pair of treatment and control participants, participants are compared by the first endpoint and where no decision for a participant with a better outcome can be reached, the comparison proceeds to the next endpoint in the hierarchy. Gehan’s scoring rule will be used for comparisons of time-to-event components [39, 40]. The component-specific pairwise comparison rules, censoring rules, and tie rules for the hierarchical composite endpoint are provided in Supplementary Table S6. This yields a number of *wins* (the treatment group participant has a better outcome in this comparison), *losses* (the control group participant has the better outcome), and *ties* (if no such conclusion can be made), where *wins* + *losses* + *ties* = *N*_pairs_ with *N*_pairs_ the number of all possible pairs of participants.

Treatment effect estimates will be presented as the Net treatment benefit (NTB): 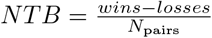

Inference will be based on asymptotic results [40, 41], primarily for reasons of computation time. The null hypothesis is *H*_0_ : *NTB* ≤ 0.

#### Common implementation rules

All methods are fit separately within each DGM, sample size, treatment-effect setting, and analysis horizon. For every fitted model, we store the estimated treatment effect, standard error, test statistic, p-value, and a model-status flag (success/non-convergence/failure). Non-converged or failed fits are summarized explicitly as part of method performance.

### Performance measures used for evaluating the method (P)

For each combination of data-generating mechanism (DGM), sample size, treatment effect scenario, analysis horizon *H* ∈ {28, 60}, and analysis method, we evaluate operating characteristics across Monte Carlo simulation replicates.

Let ***n***_*sim*_ denote the number of simulation iterations and let ***S***_*s*_ indicate whether the *s*-th simulated trial declares success according to the method-specific decision rule defined in Section Estimands and other Targets (E).

#### Type I Error and power

The rejection probability is estimated by

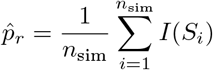

This corresponds to the proportion of simulated trials in which the null hypothesis is rejected at nominal one-sided level *α* = 0.025 in favour of the treatment. Under scenarios where the true treatment effect is null (i.e., the data-generating parameter corresponding to treatment equals zero), 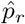 is the Type I error rate. Under alternative scenarios where a non-zero treatment effect is present, 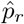 is the empirical power.

The Monte Carlo standard error (MCSE) of 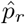 is estimated as

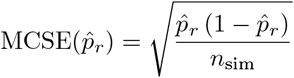

Two-sided 95% Monte Carlo confidence intervals (MC-CIs) for 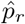 are calculated using the normal approximation:

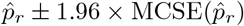

As primary analysis, operating characteristics (Type I error and power) are calculated using all simulation replicates. For these analyses, repetitions that failed to yield valid parameter estimates will be considered failures to declare success of the trial if the statistical power is estimated, and considered successes if Type I error rate is estimated (worst-case imputation). Additionally, both ways of counting failed repetitions are reported (e.g., also treating failed repetitions as successes when estimating statistical power) to provide margins of uncertainty in the estimates from simulations due to failed repetitions.

Secondary analyses estimate conditional Type I error and power using all simulation replicates for which the analysis method successfully converged. That is, inference is based on the subset of simulated trials where valid parameter estimates were obtained. Additionally, we will report the average rejection probability and the average difference to the best performing analysis approach across subsets of alternative scenarios, e.g., where a treatment effect is present.

Convergence failures are summarized separately as described in Section Technical considerations. If a substantial proportion of simulation replicates (> 1% for estimates of power, 0.1% for type I error) fails to converge for a given method and scenario, this will be investigated further, and potential causes such as model misspecification or numerical instability will be explored. Measures to mitigate failures will be taken such as switching to different packages or algorithms, increasing the maximum number of iterations in the estimation process, or introducing penalty terms for MOST.

## Overview of scenarios

The baseline and sensitivity simulation scenarios are summarized in Table 7 and specified in detail in Supplementary Table S3. The baseline full-factorial evaluation comprises 8 combinations: 2 follow-up horizons (28 and 60 days), and 4 treatment-effect settings (0, 0.75, 1.25, and 2.0 days difference in days alive and out of hospital by day *H* = 28). For scenarios analysed at H = 60, the same calibrated treatment parameter is used unless explicitly stated; the achieved DOH difference through both day 28 and day 60 will be reported from the superpopulation. The latent Brownian-motion DGM, ACTT-2 calibration setting, total sample size of 800, and 6 ordinal states are held fixed. We use the latent Brownian-motion DGM as the primary DGM because it generates clinically plausible longitudinal trajectories and is not identical to the formal analysis model of any single compared method. We have chosen a total sample size of 800 participants as base scenario because, based on clinical experience, it constitutes a feasible upper limit of a large multicentre pathogen specific trial in an inter-pandemic time. The effect sizes are chosen to reflect a null scenario, previously observed effect sizes of ARVI treatments [16, 29] and a more optimistic scenario.

**Table 7.** Parameters used in the scenarios for simulations and their specification. Parameters in the baseline scenario will be investigated in a full-factorial design (8 combinations). The additional scenarios vary sample size, scale granularity and data generating mechanisms. The default value in bold will be held constant if one of the other parameters is altered. DSF (e) = estimable Disease Specific Features, DSF (a) assumed DSF. DC = Design choices, and DC (c) = constrained DC.

| Parameter | Type | Investigated values | Description |
| --- | --- | --- | --- |
| Baseline scenario (full factorial evaluation) |  |  |  |
| Individual follow-up time | DC | 28 days, 60 days | Fixed follow-up per participant |
| Data generating mechanism (DGM) | DSF (a) | Latent Brownian motion | Other DGMs will be investigated in a subset of scenarios only. |
| DGM setting | DSF (e) | ACTT-2 (moderately ill participants) | DGMs will be calibrated to mimic previous trial's setting (see Section Overview of scenarios) |
| Total sample size | DC | 800 participants | Using a 1:1 allocation ratio |
| Number of states | DC | 6 | See Table 3 |
| Effect sizes | DSF (e) | 0, 0.75, 1.25, and 2 days | Between-group difference in time alive and out of hospital until day $H = 28$ |
| Sample size and scale granularity |  |  |  |
| Individual follow-up time | DC | 28 days, 60 days | Fixed follow-up per participant |
| Data generating mechanism (DGM) | DSF (a) | Latent Brownian motion |  |
| DGM setting | DSF (e) | ACTT-2 (moderately ill participants) | DGMs will be calibrated to mimic previous trial's setting (see Section Overview of scenarios) |
| Number of states | DC | 4, **6**, 8 | If sample sizes altered, 6 is the default |
| Total sample size | DC | 200, 400, 600, **800**, 1000 participants | Using a 1:1 allocation ratio. If scale granularity is altered, 800 is the default |
| Effect sizes | DSF (e) | 1.25 days | Between-group difference in time alive and out of hospital until $H = 28$ days |
| Data generating mechanisms |  |  |  |
| Individual follow-up time | DC | 28 days | Fixed follow-up per participant |
| Data generating mechanism (DGM) | DSF (a) | Latent Brownian motion, Longitudinal Markov model, Piecewise exponential recurrent event times with frailty | ACTT-1 and ACTT-2 settings. |
| DGM setting | DSF (e) | ACTT-1 (severely ill participants), ACTT-2 (moderately ill participants) | DGMs will be calibrated to mimic previous trial's setting (see Section Overview of scenarios) |
| Total sample size | DC | 800 participants | Using a 1:1 allocation ratio |
| Number of states | DC | 6 | See Table 3 |
| Effect sizes | DSF (e) | 0, 2 days | Between-group difference in time alive and out of hospital |
| Resampling from ACTT-2 data |  |  |  |
| Individual follow-up time | DC | 28 days | Fixed follow-up per participant |
| Data generating mechanism (DGM) | DSF (a) | ACTT-2 empirical resampling | See Section Data Generating Mechanisms (D) |
| DGM setting | DSF (e) | ACTT-2 empirical control arm for the null; ACTT-2 empirical treatment and control arms for the observed-effect scenario | DGMs will be calibrated to mimic ACTT-2 setting (see Section Overview of scenarios) |
| Total sample size | DC | 600, 800, 1000 participants | Using a 1:1 allocation ratio |
| Number of states | DC | 8 | See Table 3 |
| Effect sizes | DSF (e) | 0 days; observed ACTT-2 treatment effect | Between-group difference in time alive and out of hospital until day $H = 28$ |
| Different treatment effects |  |  |  |
| Individual follow-up time | DC | 28 days | Fixed follow-up per participant |
| Data generating mechanism (DGM) | DSF (a) | Latent Brownian motion | Using threshold specific treatment effects |
| DGM setting | DSF (e) | ACTT-1 (severely ill participants), ACTT-2 (moderately ill participants), and COV-BARRIER (moderately ill participants, higher mortality) | DGMs will be calibrated to mimic previous trial's setting (see Section Overview of scenarios and Supplementary Table S2) |
| Total sample size | DC | 800 participants | Using a 1:1 allocation ratio |
| Number of states | DC | 8 | See Table 3 |
| Effect sizes | DSF (e) | ACTT-1, ACTT-2 and COV-BARRIER state probability target values | Between-group difference not measured solely in time alive and out of hospital but state probabilities at day 28; see Supplementary Table S2 |

Null scenarios are scenarios where there is no difference in distributions of participant states between treatment groups. This implies no true difference in all derived data representations (event proportions, event times, states at time H etc.).

A more detailed investigation of, e.g., the influence of sample size and different number of states on power will be investigated in a subset of scenarios:

- varying follow-up of 28 and 60 days,
- an effect size of 1.25 days between-group difference of the time alive and out of hospital until day *H* = 28
- using the latent Brownian motion DGM

In these scenarios, dependence of power on the sample size of 200, 400, 600, 800, 1000 participants will be evaluated. Also in these scenarios, using different granularity of the ordinal scale (four and eight categories in addition to six) will be evaluated.

By default, treatment effects will be simulated using the latent Brownian motion DGM and applying a common drift across thresholds (see Section Data Generating Mechanisms (D)) to reach the desired between-group difference of the time alive and out of hospital. Additional scenarios will be investigated, where threshold-specific treatment effects are used to closely mimic ACTT-1, ACTT-2, and COV-BARRIER [33] proportions in states of treatment and control group at day 28 (target parameters are given in Supplementary Table S2).

The remaining DGMs (Ordinal Markov process and Latent event times) will be simulated for effect sizes of zero and two days difference in time alive and out of hospital and *H* = 28 days follow-up. Under each of the three mechanisms we generate data using two settings: parameters are calibrated so that the first setting matches data from ACTT-1, which included severely ill participants, and the second scenario matches ACTT-2, which included moderately ill participants. The calibration aims to mimic the baseline state probabilities, the state probabilities at day 14 and at the end of follow-up in the respective study and the proportion of participants experiencing re-hospitalisations in the control group. Calibration targets are given in Supplementary Table S2 and achieved calibration will be documented there.

Resampling from the ACTT-2 data will look at total sample sizes of 600, 800, and 1000 (1:1 allocation ratio) using the original 8-state scale and follow-up of 28 days. Effect sizes of 0 as well as the originally observed treatment effect will be investigated.

Any deviations from the pre-specified scenarios will be noted along with the reporting of results.

## Technical considerations

### Number of Simulation Iterations

The number of simulation iterations *n*_sim_ is chosen to ensure adequate Monte Carlo precision for estimating Type I error and power.

For a binomial proportion, the maximum MCSE occurs at *p*_*r*_ = 0.5, while for a one-sided Type I error close to 0.025, the MCSE is smaller. In this study, we are interested in detecting an absolute Type I error inflation of 0.5 percentage points (i.e., an increase from 0.025 to 0.030). To achieve sufficient precision relative to this target, we aim for a Monte Carlo standard error (MCSE) that is small compared to this difference.

Specifically:

- For primary Type I error scenarios, we target an MCSE of approximately 0.0025 (0.25 percentage points).
- For power scenarios (anticipated between 0.70 and 0.90), we target an MCSE ≤ 0.01.

Using

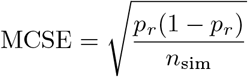

and assuming *p*_*r*_ = 0.025, achieving MCSE = 0.0025 requires approximately

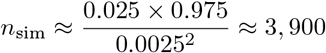

To be conservative and ensure stable estimates across all scenarios, we use at least *n*_sim_ = 5, 000 simulation iterations per scenario for the evaluation of Type I error. The same number is used for power scenarios to ensure consistent precision across performance measures.

With *n*_sim_ = 5, 000, the MCSE for a true Type I error of 0.025 is approximately 0.0022, corresponding to a 95% MC-CI half-width of about 0.0043 (i.e., total width *≈* 0.0086). This level of precision allows to detect absolute Type I error deviations of approximately 0.5 percentage points.

### Reproducibility

All simulations will set random seeds at the beginning of simulations and save the random seed at the end of each scenario. The same simulated datasets will be analysed by all methods within a scenario to allow paired comparisons across methods. The scenario identifier, R version, package versions, operating system, and full sessionInfo() output, seed stream or start and end seed, software version, and model-status flag will be stored with each simulation output.

### Convergence Failure Rate

For some analysis methods, model fitting may fail or the numerical optimization procedure may not converge for a given simulated dataset. As described in Section Common implementation rules, each fitted model is assigned a model-status flag indicating whether model fitting succeeded or resulted in failure or non-convergence.

Let *f*_*s*_ denote an indicator that the analysis method failed to converge in the *s*-th simulation replicate:

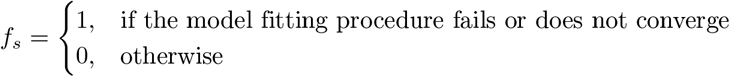

Across simulation replicates, the *failure rate* is estimated as

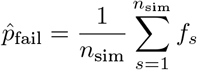

This quantity represents the proportion of simulated trials in which the analysis method fails to produce valid parameter estimates due to numerical failure or lack of convergence.

The Monte Carlo standard error (MCSE) of 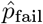 is estimated using the binomial approximation

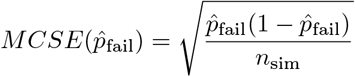

Two-sided 95% Monte Carlo confidence intervals are calculated as

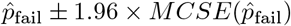

Convergence failure rates will be reported for each combination of data-generating mechanism, sample size, treatment effect scenario, analysis horizon, and analysis method. These results provide an assessment of the numerical stability and practical feasibility of each analysis approach under the simulated trial conditions.

If an analysis strategy proves infeasible due to convergence issues or other types of failures, in certain scenarios, an alternative analysis strategy is sought. These deviations from the protocol and the resulting findings will then be reported in detail.

### Software

We use R (version 4.5 or higher) for all analyses, including standard logistic regression models and t-tests [42]. MOST and proportional odds models will be fit using the VGAM package [43] (version 1.1-14 or higher) or rms package [44] (version 8.1-1 or higher), cluster robust standard errors for VGAM objects will be calculated using the markov.misc package [45]. Time to event models will be fit using the survival package [46] (Version 3.8-6 or higher). Generalized pairwise comparisons will use the BuyseTest package [47] (Version 3.3.9 or higher).

## Final Remarks

This simulation study investigates the statistical power and Type I error rates of several endpoint definitions and associated analysis approaches for the primary analysis of clinical trials, specifically in the field of ARVI. Our results will aid the choice of analysis approaches in future trials.

The operating characteristics investigated in our simulations will be summarized by mean and uncertainty quantified by suitable Monte Carlo standard errors.

We compare different endpoint definitions and corresponding analysis methods that do not target the same estimand. Provided that Type I error is adequately controlled, empirical power will provide one important component of the assessment of whether an endpoint-analysis strategy is feasible as a primary analysis, alongside interpretability, clinical relevance, robustness, and handling of intercurrent events.

Several extensions are of interest for future work. First, while our choice of data generating mechanisms aims at testing very different assumptions about the way the data were simulated they do not systematically evaluate violations of certain assumptions made in newer analysis approaches. Understanding operating characteristics under specific violation of assumptions will be important.

### Limitations

The handling of missing data and loss to follow-up are not unambiguous for the methods compared in this paper and thus difficult to compare. Drop-out and associated choices could have an impact on the performance of the different approaches. Such differences may be aggravated when longer follow-up periods are used (e.g., 60 days). However, such comparisons are beyond the scope of our study.

The proposed simulation study only considers a frequentist framework. Although there has been some work using Bayesian longitudinal ordinal models [16], systematic neutral comparison studies are lacking and could be an interesting goal for future research.

The data that we use for calibration of the DGM stems from trials in populations with COVID-19. Different ARVI, such as RSV and influenza, likely have different distributions of participant states over time, which could affect operating characteristics of analysis approaches.

Finally, the data generating mechanisms we consider are not designed to provide mechanistic insight into potential performance differences. Systematic investigations into time-varying treatment effects, or how treatment effects for only some state transitions impact power might be interesting for future studies.

## Results Dissemination

Results from this simulation study will be disseminated through open access, peer-reviewed journals and presentations at relevant conferences.

Within the scope of the PROACT EU-RESPONSE project, preliminary sample size calculations were conducted using a single data generating mechanism that resembles the latent brownian motion DGM described in Section Data Generating Mechanisms (D). Previous simulations are stored and time-stamped in a public Git repository (github.com/scjohannes/proact-eu-response). The simulation functions developed so far are publicly available (github.com/scjohannes/markov.misc).

The simulation code, analysis code, and scenario specifications will be made publicly available in a public repository upon publication. The ACTT-2 individual participant data used for calibration/resampling are not generated by the authors and are subject to third-party access controls and applicable data-use restrictions. Requests for access to the underlying ACTT-2 data can be filed through the NIH data portal (https://accessclinicaldata.niaid.nih.gov/).

## Data Availability

Previous simulations are stored and time-stamped in a public Git repository (github.com/scjohannes/proact-eu-response). The simulation functions developed so far are publicly available (github.com/scjohannes/markov.misc). The simulation code, analysis code, and scenario specifications will be made publicly available in a public repository upon publication. The ACTT-2 individual participant data used for calibration/resampling are not generated by the authors and are subject to third-party access controls and applicable data-use restrictions. Requests for access to the underlying ACTT-2 data can be filed through the NIH data portal (https://accessclinicaldata.niaid.nih.gov/).

https://github.com/scjohannes/markov.misc

https://accessclinicaldata.niaid.nih.gov/

## Acknowledgements

We acknowledge the US National Institute of Allergy and Infectious Disease Clinical Trials Data Repository with regards to the ACTT-2 trial data. We acknowledge the use of large language models as coding and writing aids.

